# Hidden in Plain Sight: Clinical Profile, Diagnosis, and Outcomes of Cardiac Amyloidosis in Cali, Colombia

**DOI:** 10.64898/2026.10.15.26365704

**Authors:** Stephany Barbosa Balaguera, Orlando Castaño, Eduardo José Echeverry, Christian David Messu Llano, Álvaro José Muriel Ruiz, Jacobo Barona Díaz, Ángela María Timarán Córdoba, Álvaro Herrera Escandón

## Abstract

**Background:** Cardiac amyloidosis is an increasingly recognized cause of heart failure in older adults, yet data from Colombia and, in particular, from the country’s southwest region remain scarce. We describe the clinical, diagnostic, imaging, and outcome profile of patients with cardiac amyloidosis at two high-complexity institutions in Cali.

**Methods:** Observational, descriptive, retrospective, two-center study. Patients were identified through the institutional registries of both centers using ICD-10 diagnostic codes for amyloidosis, and clinical information was collected manually from medical records. Continuous variables are reported as median (interquartile range) and categorical variables as counts and percentages, with the number of patients with available data specified for each variable.

**Results:** Thirty-two patients were analyzed (median age 77 years; 68.8% men). Heart failure was nearly universal (90.6%) and atrial fibrillation was frequent (46.9%); carpal tunnel syndrome was present in 31.2%. Bone scintigraphy showed positive myocardial uptake (Perugini 2–3) in 88.5%, and the Val142Ile variant was found in all genotyped patients, consistent with a predominance of transthyretin amyloidosis. Left ventricular ejection fraction was preserved in most patients despite marked concentric hypertrophy. Tafamidis was used in 37.5%. Overall mortality was 35.5%, and NT-proBNP was markedly higher among patients who died (median 6640 vs 1186 pg/mL; p=0.003).

**Conclusions:** In this southwest Colombian cohort, cardiac amyloidosis presented as the classic transthyretin phenotype of the older male with hypertrophic, non-dilated heart failure, a high yield of non-invasive diagnosis, and a striking predominance of the Val142Ile variant. Mortality was high and closely linked to NT-proBNP.

## Introduction

Cardiac amyloidosis results from the extracellular deposition of misfolded proteins in the myocardium, producing a progressive, infiltrative and typically restrictive cardiomyopathy. Long regarded as a rare disease, it is now recognized as an under-diagnosed and increasingly important cause of heart failure in older adults.^1,2,20,21^ Two proteins account for the overwhelming majority of cases: transthyretin (ATTR), which may be wild-type or hereditary, and immunoglobulin light chains (AL).^2,3^ Distinguishing between them is essential because their treatment and prognosis differ substantially.^3,4^ The true burden of the disease has been consistently underestimated. Screening studies suggest that cardiac amyloidosis is present in a meaningful proportion of patients with heart failure with preserved ejection fraction, aortic stenosis in the elderly, and unexplained left ventricular hypertrophy.^5,6^ The turning point in diagnosis was the validation of bone scintigraphy, which allows a reliable, non-invasive diagnosis of ATTR cardiac amyloidosis once a monoclonal protein has been excluded, avoiding endomyocardial biopsy in most cases.^2,7,9,10^ In parallel, disease-modifying therapy — in particular the transthyretin stabilizer tafamidis — has changed the outlook for ATTR, making timely diagnosis clinically consequential.^11,12,13^

These advances have not been distributed evenly. In Latin America, awareness remains limited and access to scintigraphy and to disease-modifying therapy is uneven, which translates into persistent underdiagnosis and diagnostic delay.^14^ Colombian data are still scarce and come from only a handful of referral centers,^15,16^ and evidence from the country’s southwest is virtually absent. We therefore aimed to describe the clinical, diagnostic, imaging, and outcome profile of patients with cardiac amyloidosis managed at two high-complexity institutions in Cali, in order to inform local suspicion, diagnosis, and management.

## Methods

### Study design and setting

This was an observational, descriptive, retrospective, two-center study conducted at two high-complexity institutions in Cali, southwestern Colombia: Clínica Imbanaco and Clínica Sebastián de Belalcázar.

### Population and case identification

Patients with a diagnosis of cardiac amyloidosis were identified through the institutional registries of both participating centers using ICD-10 diagnostic codes for amyloidosis (E85). Clinical information was collected manually from the electronic and physical medical records by the investigators. Patients with complete clinical records were included in the analysis; when a specific variable was not documented, the corresponding patient was excluded only from the analysis of that variable, and the number of patients with available data is reported throughout.

### Variables and definitions

Collected variables included demographics, cardiovascular comorbidities, symptoms and New York Heart Association (NYHA) functional class, extracardiac “red flags” (carpal tunnel syndrome, spinal canal stenosis, biceps tendon rupture), electrocardiography, transthoracic echocardiography with global longitudinal strain, cardiac magnetic resonance, ambulatory blood-pressure and Holter monitoring, biomarkers, 99mTc-pyrophosphate (PYP) scintigraphy graded with the Perugini scale^10^, genetic testing, serum and urine monoclonal studies, treatment, and vital status. Relative wall thickness and left ventricular mass (Devereux formula) were derived from echocardiographic measurements. Amyloidosis subtype was approached following the accepted non-invasive algorithm^9^ (positive scintigraphy with exclusion of a monoclonal component for ATTR, and the presence of a monoclonal component for AL); because a single validated subtype variable was not available for every patient, the subtype distribution is presented as provisional.

### Statistical analysis

Continuous variables are summarized as median (interquartile range, IQR) and categorical variables as absolute frequencies and percentages calculated over the number of patients with available data. For the exploratory comparison between patients who died and those who survived, the Mann-Whitney U test was used for continuous variables and the Fisher exact test for categorical variables; correlations were assessed with the Spearman coefficient. A two-sided p value below 0.05 was considered statistically significant. Analyses were performed in Python (pandas and SciPy).

### Ethical considerations

The study was approved by the Research Ethics Committee of Clínica Sebastián de Belalcázar and was based on the retrospective review of clinical records. Patient information was handled in an anonymized manner, with identifiers removed and replaced by a study code for analysis.

## Results

### Baseline characteristics

Thirty-two patients were analyzed. The cohort consisted of older adults with a clear male predominance (median age 77 years [IQR 72.8–83.2]; 22 men, 68.8%). Heart failure was present in almost all patients (29 (90.6%)) and atrial fibrillation in nearly half (15 (46.9%)). Hypertension (19 (59.4%)), chronic kidney disease (12 (37.5%)) and dyslipidemia (12 (37.5%)) were common. Dyspnea was the dominant symptom (17 (53.1%)), with most patients in NYHA class II. Among the extracardiac red flags, carpal tunnel syndrome stood out, present in 10 (31.2%) of patients. Baseline characteristics are shown in Table 1 and Figure 1.

**Table 1.** Baseline clinical, laboratory, and echocardiographic characteristics of the cohort (n=32). IQR, interquartile range; LVEF, left ventricular ejection fraction; TAPSE, tricuspid annular plane systolic excursion; PASP, pulmonary artery systolic pressure; LV, left ventricular.

| Characteristic | Value | n |
| --- | --- | --- |
| **Demographics** |  |  |
| Age, years — median (IQR) | 77 (72.8–83.2) | 32 |
| Male sex, n (%) | 22 (68.8%) | 32 |
| **Comorbidities, n (%)** |  |  |
| Heart failure | 29 (90.6%) | 32 |
| Hypertension | 19 (59.4%) | 32 |
| Atrial fibrillation | 15 (46.9%) | 32 |
| Dyslipidemia | 12 (37.5%) | 32 |
| Chronic kidney disease | 12 (37.5%) | 32 |
| Hypothyroidism | 11 (34.4%) | 32 |
| Diabetes mellitus | 8 (25%) | 32 |
| Coronary artery disease | 7 (21.9%) | 32 |
| Former smoking | 12 (38.7%) | 31 |
| **Symptoms and functional class, n (%)** |  |  |
| Dyspnea | 17 (53.1%) | 32 |
| Palpitations | 5 (15.6%) | 32 |
| Chest pain | 4 (12.5%) | 32 |
| Syncope | 2 (6.2%) | 32 |
| NYHA class III–IV | 6 (19.4%) | 31 |
| **Extracardiac red flags, n (%)** |  |  |
| Carpal tunnel syndrome | 10 (31.2%) | 32 |
| Spinal canal stenosis | 3 (9.4%) | 32 |
| **Laboratory — median (IQR)** |  |  |
| NT-proBNP, pg/mL | 2342 (902–7761.5) | 27 |
| Troponin | 31.6 (26.5–67.5) | 15 |
| Creatinine, mg/dL | 1.1 (1–1.4) | 29 |
| Hemoglobin, g/dL | 14 (12.9–14.7) | 24 |
| **Echocardiography — median (IQR)** |  |  |
| Septal wall thickness, mm | 16 (15–18) | 30 |
| Posterior wall thickness, mm | 15 (13–16) | 28 |
| LVEF, % | 55 (45–60) | 31 |
| Left atrium, mm | 48 (41–55) | 25 |
| TAPSE, mm | 18 (15.5–19.5) | 27 |
| PASP, mmHg | 40 (33–42) | 25 |
| Concentric hypertrophy, n (%) | 28 (96.6%) | 29 |
| Relative wall thickness, median | 0.7 | 27 |
| LV mass (Devereux), g — median (IQR) | 271 (213–318) | 27 |
| Preserved LVEF ($\geq 50\%$), n (%) | 22 (71%) | 31 |
| Pericardial effusion, n (%) | 7 (24.1%) | 29 |

**Figure 1.**
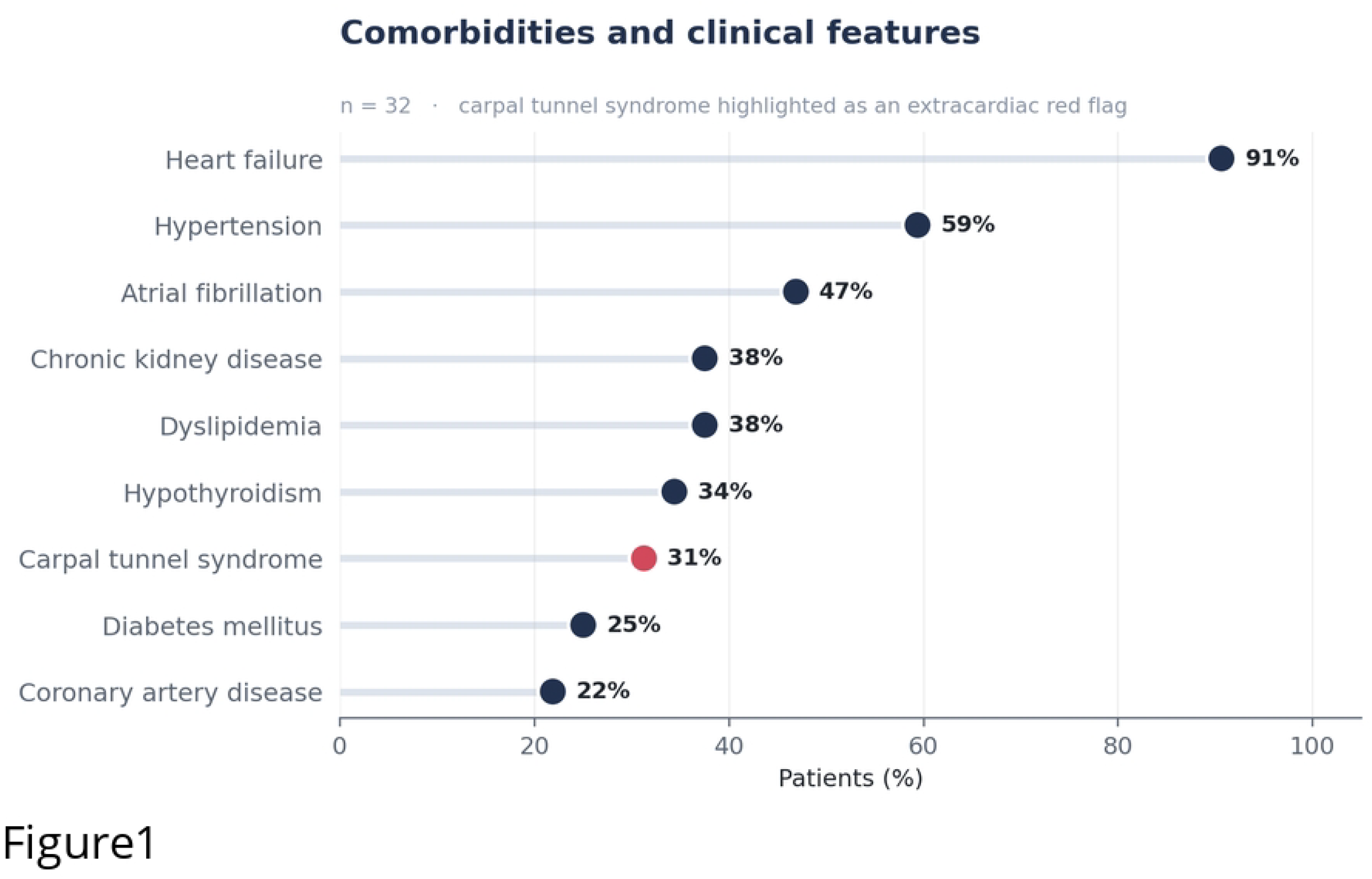
Comorbidities and clinical features. Carpal tunnel syndrome, an extracardiac red flag, is highlighted.

On echocardiography, the myocardium was markedly thickened (median septal thickness 16 mm) with a concentric, non-dilated pattern; concentric hypertrophy was present in 28 (96.6%) of assessable studies and the median relative wall thickness was 0.7, with an elevated estimated left ventricular mass (median 271 g). Despite this degree of infiltration, the left ventricular ejection fraction was preserved (≥50%) in 22 of 31 patients (71%), reflecting a predominantly hypertrophic, heart-failure-with-preserved-ejection-fraction phenotype. Global longitudinal strain was reduced (median −12%), and relative apical sparing was explicitly described in 2 of the 16 studies reporting strain. NT-proBNP was elevated (median 2342 pg/mL [IQR 902– 7761.5]).

### Diagnostic work-up

Bone scintigraphy with 99mTc-PYP showed positive myocardial uptake (Perugini grade 2 or 3) in 23 (88.5%) of studied patients, with grade 3 in 15 (Figure 2). Genetic testing was positive in 9 of 18 patients tested, and — most notably — the Val142Ile (p.Val142Ile) transthyretin variant was identified in every genotyped patient (8/8). A monoclonal component on immunofixation was detected in 4/19 of those studied. Taken together, these findings indicate a predominance of transthyretin amyloidosis; the provisional subtype distribution is shown in Table 2 and Figure 3.

**Table 2.** Diagnostic findings and provisional etiologic subtype. PYP, 99mTc-pyrophosphate; ATTR, transthyretin amyloidosis; AL, light-chain amyloidosis.

| Diagnostic finding | Value | n |
| --- | --- | --- |
| Positive PYP uptake (Perugini 2–3), n (%) | 23 (88.5%) | 26 |
| Perugini grade 3, n | 15 | 26 |
| Positive genetic test, n | 9 | 18 |
| Val142Ile variant (of genotyped), n (%) | 8 (100%) | 8 |
| Monoclonal component (immunofixation), n (%) | 4 (21%) | 19 |
| **Etiologic subtype (provisional), n** |  |  |
| ATTR (scintigraphy + genetic) | 24 | 32 |
| AL (probable) | 5 | 32 |
| Indeterminate | 1 | 32 |

**Figure 2.**
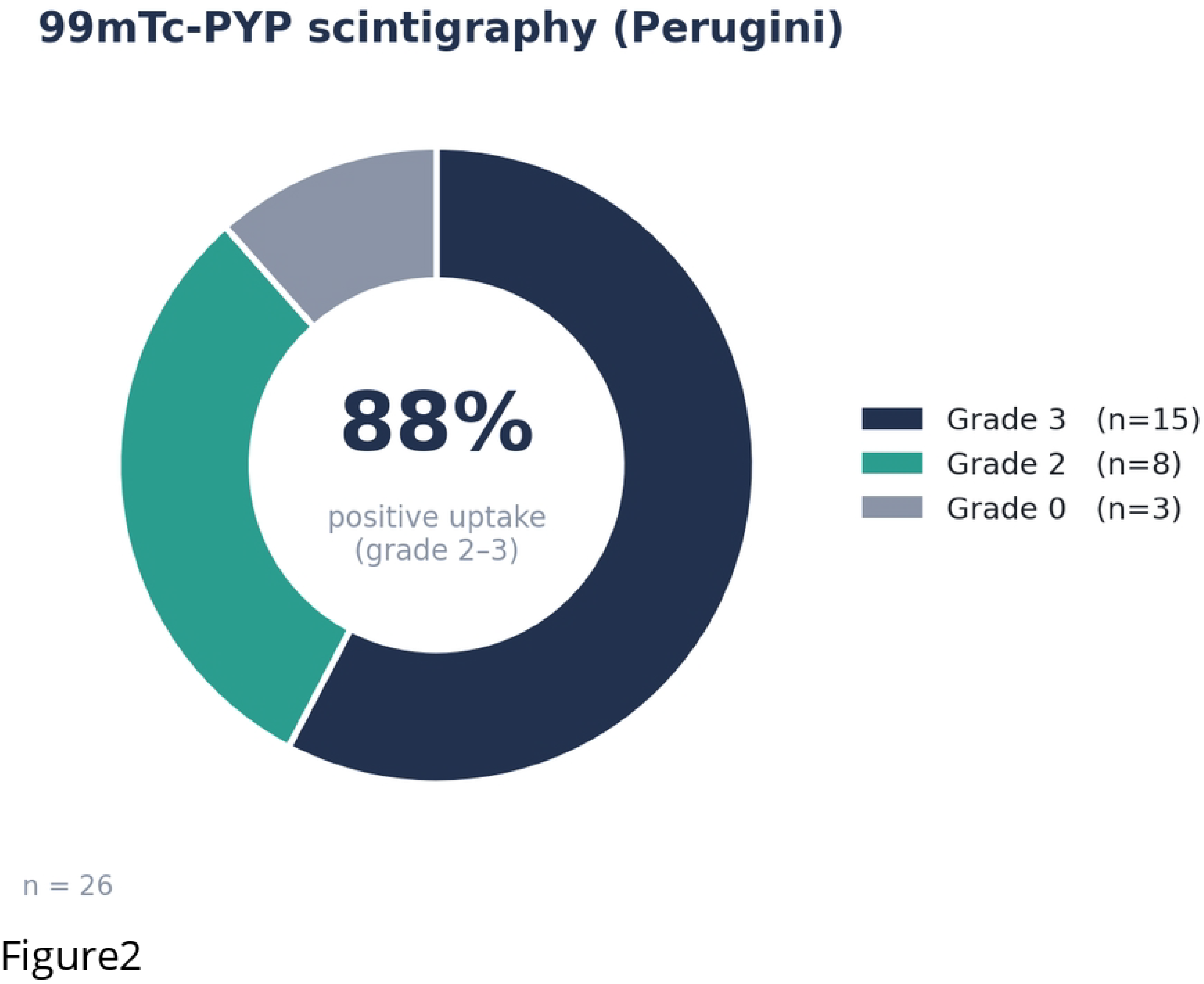
Distribution of 99mTc-PYP scintigraphy (Perugini grade).

**Figure 3.**
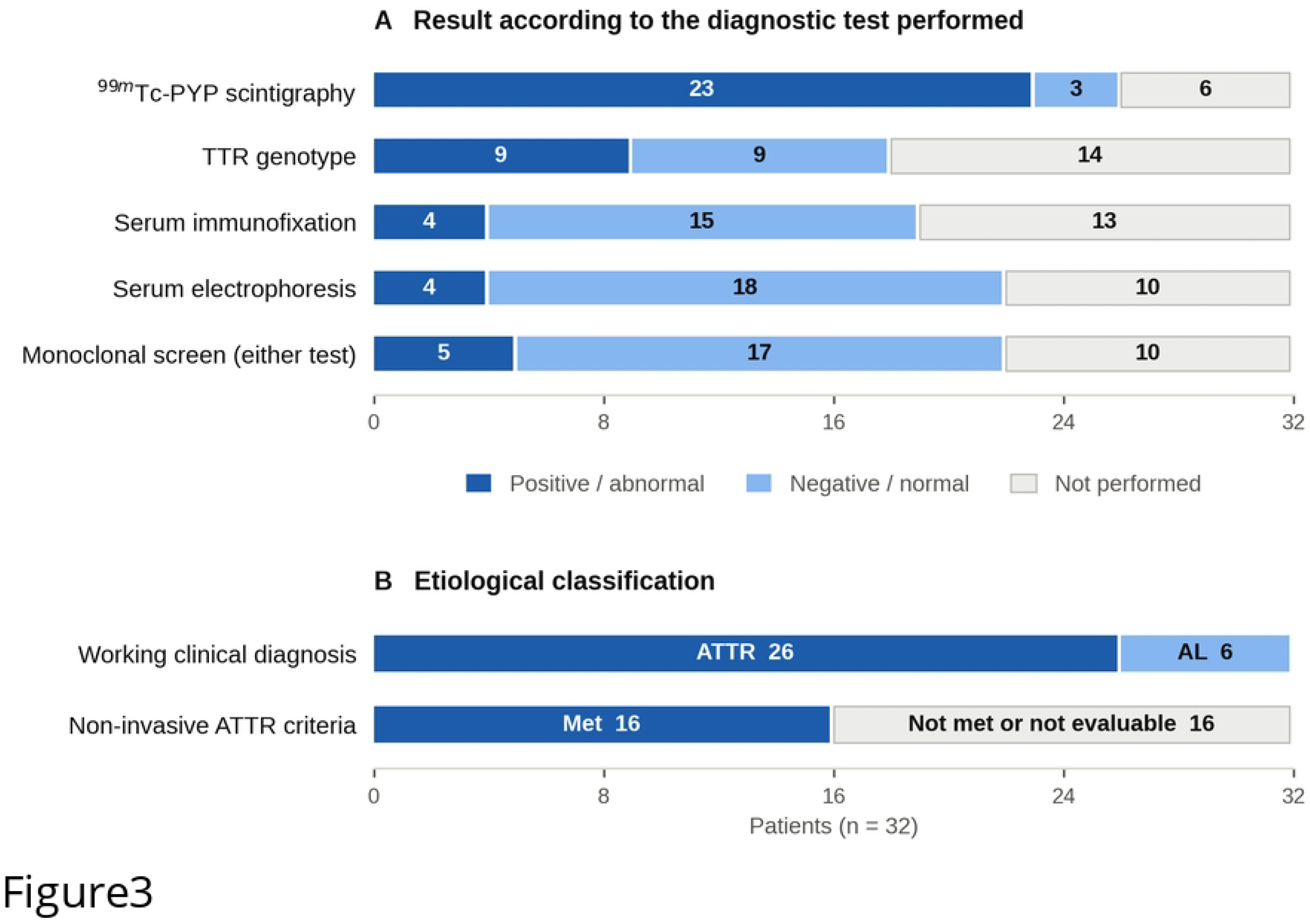
Provisional distribution of etiologic subtype. ATTRv, hereditary transthyretin amyloidosis.

### Treatment

Tafamidis was used in 12 (37.5%) of patients. Heart-failure therapy was frequent, including SGLT2 inhibitors in 27 (84.4%), diuretics in 21 (65.6%), mineralocorticoid receptor antagonists in 18 (56.2%) and beta-blockers in 17 (53.1%); approximately half of the patients were anticoagulated. Details are shown in Table 3.

**Table 3.** Pharmacological treatment. ARB, angiotensin-receptor blocker; ACE, angiotensin-converting enzyme; ARNI, angiotensin receptor-neprilysin inhibitor.

| Treatment | In use, n (%) | n |
| --- | --- | --- |
| Tafamidis | 12 (37.5%) | 32 |
| SGLT2 inhibitor | 27 (84.4%) | 32 |
| Diuretics | 21 (65.6%) | 32 |
| Mineralocorticoid receptor antagonist | 18 (56.2%) | 32 |
| Beta-blocker | 17 (53.1%) | 32 |
| Direct oral anticoagulant | 17 (53.1%) | 32 |
| ARB | 14 (43.8%) | 32 |
| Calcium-channel blocker | 13 (40.6%) | 32 |
| ACE inhibitor | 7 (21.9%) | 32 |
| ARNI | 2 (6.2%) | 32 |

### Outcomes

Overall mortality was 11 (35.5%). In the exploratory comparison between patients who died and those who survived, NT-proBNP was markedly higher among those who died (median 6640 vs 1186 pg/mL; p=0.003; Figure 4), and both septal and posterior wall thickness were greater. Tafamidis use was less frequent among patients who died; this association should be interpreted with caution (see Discussion). Results are summarized in Table 4.

**Table 4.** Exploratory comparison between deceased and surviving patients (n=31, 11 events). Continuous variables are medians. p values from Mann-Whitney U or Fisher exact test, without adjustment for multiple comparisons or confounders.

| Variable | Deceased (n=11) | Alive (n=20) | p |
| --- | --- | --- | --- |
| NT-proBNP, pg/mL (median) | 6640 | 1186 | 0.003 |
| Septal thickness, mm | 18 | 16 | 0.027 |
| Posterior wall, mm | 16 | 14 | 0.036 |
| LVEF, % | 50 | 59 | 0.132 |
| Pericardial effusion, n | 4/9 | 2/19 | 0.064 |
| Tafamidis use, n | 1/11 | 10/20 | 0.047 |

**Figure 4.**
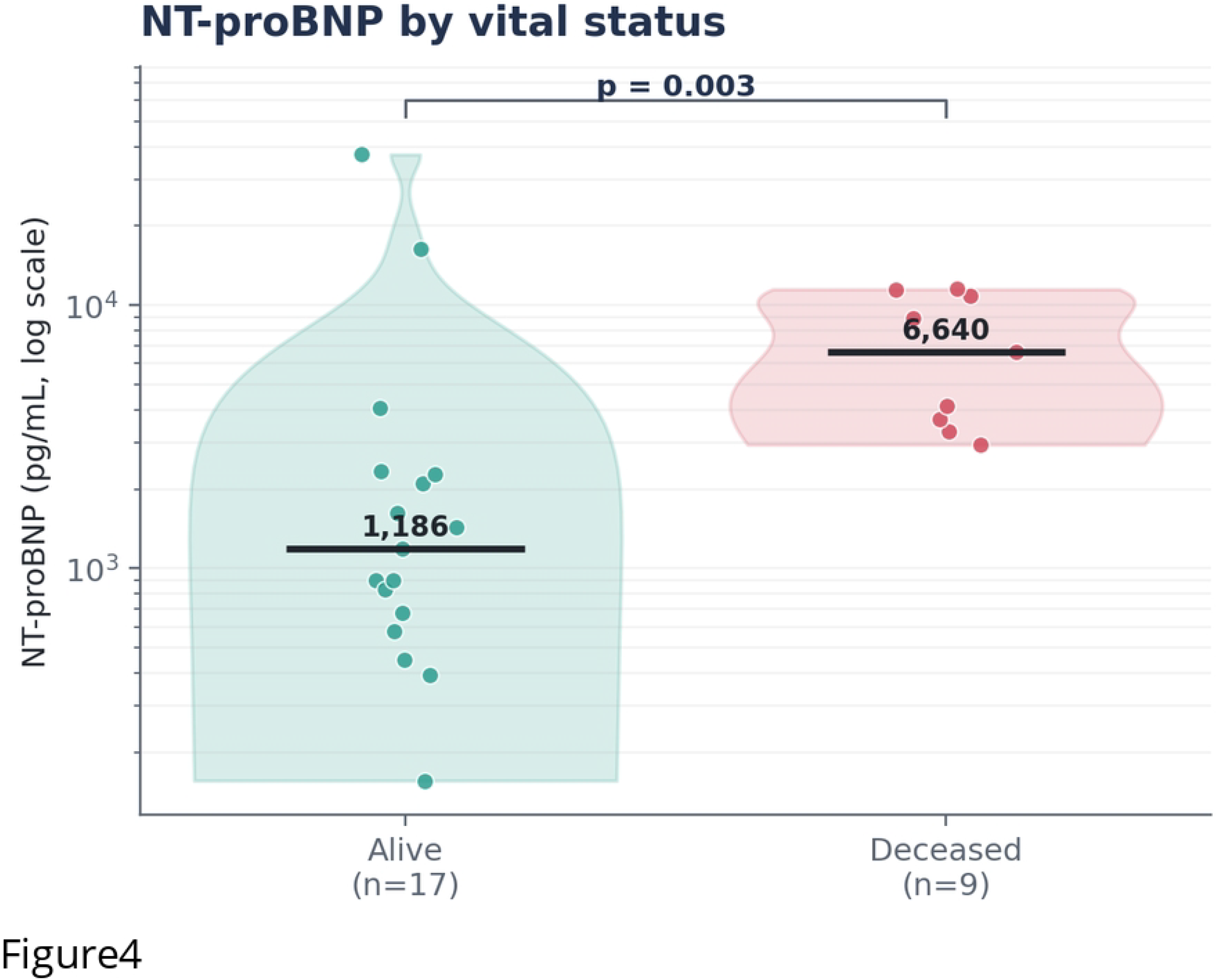
NT-proBNP according to vital status (log scale). Horizontal bars indicate medians.

## Discussion

In this two-center cohort from two high-complexity institutions in Cali, cardiac amyloidosis presented as the classic transthyretin phenotype: an older man with heart failure, a thickened and non-dilated left ventricle, frequently preserved ejection fraction, atrial fibrillation, and carpal tunnel syndrome as a frequent antecedent red flag.^1,2,17^ The high proportion of positive bone scintigraphy and the reduced global longitudinal strain with relative apical sparing are consistent with the characteristic multimodal imaging signature of the disease.^7,8,18^

The most distinctive finding was the uniform presence of the Val142Ile transthyretin variant among genotyped patients. This variant is well recognized as the most common amyloidogenic TTR mutation in individuals of African ancestry, and its predominance is biologically and epidemiologically coherent in the population of southwestern Colombia and the Pacific region. This observation reinforces the need for a low threshold for genetic testing and family screening in this setting.^2,19^

### Comparison with other cohorts

Our findings are consistent with the two previously published Colombian series. In the case series from a university hospital in Cali (14 patients), patients were also predominantly male and were diagnosed through bone scintigraphy and TTR sequencing, with the Val142Ile variant identified in all hereditary ATTR cases; no deaths had occurred at the time of that report, in keeping with its smaller size and shorter follow-up.^15^ The referral-center cohort from Bogotá (31 patients) reported a comparable median age, a similar burden of atrial fibrillation and heart failure, and an overall mortality of 25.8%, in the same range as our 35.5%; in that cohort, disease-modifying therapy for ATTR was not available during the study period.^16^ Our cohort, of comparable size, adds data from previously unreported centers in southwestern Colombia and is characterized by a marked predominance of transthyretin amyloidosis and of the Val142Ile variant. At the regional level, our experience mirrors the gaps described by the AMILO-LATAM group, namely underdiagnosis and uneven access to scintigraphy and to disease-modifying therapy across Latin America.^14^ Beyond these three references, directly comparable single-center Colombian cohorts are lacking, and this remaining gap should be acknowledged rather than filled with indirect comparisons.

### Prognosis and treatment

Mortality in our cohort was high and was strongly associated with NT-proBNP, in line with the central prognostic role of natriuretic peptides and staging systems in cardiac amyloidosis.^12,18,22^ The lower use of tafamidis among patients who died should not be interpreted as evidence of a treatment effect: in a retrospective cohort this association is very likely to reflect indication and survival bias, since patients with more advanced disease, with AL amyloidosis, or who died early may never have received the drug.^11,12,13^ The observed use of tafamidis in roughly one-third of patients, in a context where access has historically been limited across the region, is itself a relevant description of real-world care.^14^

### Strengths and limitations

The main strength of this study is its detailed, multimodal phenotyping — combining clinical, electrocardiographic, echocardiographic, strain, cardiac magnetic resonance, Holter, biomarker, scintigraphic and genetic data — in a real-world cohort from a region of Colombia not previously represented in the literature. Data were handled in an anonymized manner. The study also has important limitations. The sample size is small and the design is retrospective and limited to two centers, which limits generalizability and statistical power; consequently, the comparison between deceased and surviving patients is exploratory and hypothesis-generating, was not adjusted for multiple comparisons or confounders, and should be read as such. There was a substantial amount of missing data for several variables, so the denominator varies across analyses. A single validated variable for amyloidosis subtype was not available for every patient, so the ATTR-versus-AL distribution is provisional and should be confirmed on a case-by-case basis using the accepted non-invasive algorithm.^9^ Finally, reliable dates of diagnosis and follow-up were not available, which precluded a time-to-event (survival) analysis; incorporating these dates in future work would allow Kaplan-Meier and Cox analyses.

## Conclusion

In these two high-complexity institutions in Cali, cardiac amyloidosis presented as the classic transthyretin phenotype of the older male with hypertrophic, non-dilated heart failure, a high yield of non-invasive diagnosis, and a striking predominance of the Val142Ile variant. Mortality was high and closely linked to NT-proBNP. These findings support a higher index of suspicion, earlier use of the non-invasive diagnostic algorithm, and broader access to disease-modifying therapy for cardiac amyloidosis in southwestern Colombia.

## Data Availability

The data underlying this study cannot be made publicly available because they consist of identifiable clinical information abstracted from the medical records of a small cohort of patients with a rare disease. Ethical and legal restrictions apply. The study was approved by the Research Ethics Committee of Clínica Sebastián de Belalcázar (approval 219-26 UNV, minutes No. 030-26, 30 July 2026) on the condition that the clinical records be reviewed within the participating institutions and analysed under a study code. The committee waived the requirement for individual written informed consent on that basis, and the patients consequently never consented to the public release of their data. Public deposition is further restricted by Colombian Resolution 8430 of 1993, which governs the confidentiality of clinical records in health research, and by Colombian Law 1581 of 2012 on the protection of personal data. Even in de-identified form, the dataset carries a material risk of re-identification. It comprises only 32 patients with a rare disease, identified at two named institutions in a single city, and includes age, sex, transthyretin genotype, imaging measurements and biomarker values. A clinician practising in the region could plausibly identify individual patients from that combination of variables. A de-identified dataset is nevertheless available to qualified researchers upon reasonable request. Requests should be addressed to the Research Ethics Committee at, which evaluates each request and is the body responsible for authorizing access. The committee is independent of the authors, and no author is the sole point of contact for these data. The authors did not have special access privileges to these data that other researchers would not have upon approval by the committee.

## Supporting information

**S1 Checklist**. STROBE checklist.

**S2 File**. Graphical abstract.

## References

1. Ruberg FL, Maurer MS. Cardiac amyloidosis due to transthyretin protein: a review. JAMA. 2024;331(9):778–791.

2. Garcia-Pavia P, Rapezzi C, Adler Y, Arad M, Basso C, Brucato A, et al. Diagnosis and treatment of cardiac amyloidosis: a position statement of the ESC Working Group on Myocardial and Pericardial Diseases. Eur Heart J. 2021;42(16):1554–1568.

3. Falk RH, Alexander KM, Liao R, Dorbala S. AL (light-chain) cardiac amyloidosis: a review of diagnosis and therapy. J Am Coll Cardiol. 2016;68(12):1323–1341.

4. Gertz M. Cardiac amyloidosis. Heart Fail Clin. 2022;18(3):479–488.

5. Aimo A, Merlo M, Porcari A, Georgiopoulos G, Pagura L, Vergaro G, et al. Redefining the epidemiology of cardiac amyloidosis. A systematic review and meta-analysis of screening studies. Eur J Heart Fail. 2022;24(12):2342–2351.

6. González-López E, Gallego-Delgado M, Guzzo-Merello G, et al. Wild-type transthyretin amyloidosis as a cause of heart failure with preserved ejection fraction. Eur Heart J. 2015;36:2585–2594.

7. Dorbala S, Cuddy S, Falk RH. How to image cardiac amyloidosis: a practical approach. JACC Cardiovasc Imaging. 2020;13(6):1368–1383.

8. Ioannou A, Patel R, Gillmore JD, Fontana M. Imaging-guided treatment for cardiac amyloidosis. Curr Cardiol Rep. 2022;24(7):839–850.

9. Gillmore JD, Damy T, Fontana M, Hutchinson M, Lachmann HJ, Martinez-Naharro A, et al. Nonbiopsy diagnosis of cardiac transthyretin amyloidosis. Circulation. 2016;133:2404–2412.

10. Perugini E, Guidalotti PL, Salvi F, et al. Noninvasive etiologic diagnosis of cardiac amyloidosis using 99mTc-3,3-diphosphono-1,2-propanodicarboxylic acid scintigraphy. J Am Coll Cardiol. 2005;46:1076–1084.

11. Stern LK, Patel J. Cardiac amyloidosis treatment. Methodist Debakey Cardiovasc J. 2022;18(2):59–72.

12. Kittleson MM, Ruberg FL, Ambardekar AV, Brannagan TH, Cheng RK, Clarke JO, et al. 2023 ACC expert consensus decision pathway on comprehensive multidisciplinary care for the patient with cardiac amyloidosis. J Am Coll Cardiol. 2023;81(11):1076–1126.

13. Maurer MS, Schwartz JH, Gundapaneni B, Elliott PM, Merlini G, Waddington-Cruz M, et al. Tafamidis treatment for patients with transthyretin amyloid cardiomyopathy. N Engl J Med. 2018;379:1007–1016.

14. Mut F, Carvajal I, Camilletti J, Erriest J, Alexanderson E, Grossman GB. Cardiac amyloidosis in Latin America: gaps and opportunities to increase awareness of the disease. Findings from the AMILO-LATAM research group. J Nucl Cardiol. 2022. doi:10.1007/s12350-022-03005-5.

15. Lopez-Ponce de Leon JD, Granados-Alvarez S, Arango-Ibanez JP, Montero Echeverri JM, Arteaga Tobar AA, Florez-Garcia AF, et al. A case series of patients with cardiac amyloidosis evaluated at a Colombian university hospital. Front Cardiovasc Med. 2025;12:1487717.

16. Ortiz P, Polania-Sandoval CA, Gaviria-Villegas J, Gutierrez-Villamil C, Zuluaga-Arbelaez C, Marin-Oyaga V, et al. Clinical characterization of patients with cardiac amyloidosis in a referral center of Colombia. Arch Cardiol Mex. 2024;94(4):451–458.

17. Griffin JM, Rosenblum H, Maurer MS. Pathophysiology and therapeutic approaches to cardiac amyloidosis. Circ Res. 2021;128(10):1554–1575.

18. Fontana M, Ioannou A, Cuddy S, Dorbala S, Masri A, Moon JC, et al. The last decade in cardiac amyloidosis: advances in understanding pathophysiology, diagnosis and quantification, prognosis, treatment strategies, and monitoring response. JACC Cardiovasc Imaging. 2025;18(4):478–499.

19. Danilov A, D’Angelo L, Marsela E, Costabel JP, Jorde UP, Rochlani Y. Transthyretin cardiac amyloidosis: advances and ambiguities. Heart Fail Rev. 2025;30(6):1341–1352.

20. Martinez-Naharro A, Hawkins PN, Fontana M. Cardiac amyloidosis. Clin Med (Lond). 2018;18(Suppl 2):s30–s35.

21. Laptseva N, Benz DC, Schwotzer R, Flammer AJ. Cardiac amyloidosis. Swiss Med Wkly. 2024;154:4186.

22. Grogan M, Scott CG, Kyle RA, et al. Natural history of wild-type transthyretin cardiac amyloidosis and risk stratification using a novel staging system. J Am Coll Cardiol. 2016;68:1014–1020.

